# Enhancing Brain Age Estimation Under Uncertainty: A Spectral-normalized Neural Gaussian Process Approach Utilizing 2.5D Slicing

**DOI:** 10.1101/2025.02.26.25322983

**Authors:** Zeqiang Linli, Xingcheng Liang, Zhenhua Zhang, Kang Hu, Shuixia Guo

## Abstract

Brain age gap, the difference between estimated brain age and chronological age via magnetic resonance imaging, has emerged as a pivotal biomarker in the detection of brain abnormalities. While deep learning is accurate in estimating brain age, the absence of uncertainty estimation may pose risks in clinical use. Moreover, current 3D brain age models are intricate, and using 2D slices hinders comprehensive dimensional data integration. Here, we introduced Spectral-normalized Neural Gaussian Process (SNGP) accompanied by 2.5D slice approach for seamless uncertainty integration in a single network with low computational expenses, and extra dimensional data integration without added model complexity. Subsequently, we compared different deep learning methods for estimating brain age uncertainty via the Pearson correlation coefficient, a metric that helps circumvent systematic underestimation of uncertainty during training. SNGP shows excellent uncertainty estimation and generalization on a dataset of 11 public datasets (N=6327), with competitive predictive performance (MAE=2.95). Besides, SNGP demonstrates superior generalization performance (MAE=3.47) on an independent validation set (N=301). Additionally, we conducted five controlled experiments to validate our method. Firstly, uncertainty adjustment in brain age estimation improved the detection of accelerated brain aging in adolescents with ADHD, with a 38% increase in effect size after adjustment. Secondly, the SNGP model exhibited OOD detection capabilities, showing significant differences in uncertainty across Asian and non-Asian datasets. Thirdly, the performance of DenseNet as a backbone for SNGP was slightly better than ResNeXt, attributed to DenseNet’s feature reuse capability, with robust generalization on an independent validation set. Fourthly, site effect harmonization led to a decline in model performance, consistent with previous studies. Finally, the 2.5D slice approach significantly outperformed 2D methods, improving model performance without increasing network complexity. In conclusion, we present a cost-effective method for estimating brain age with uncertainty, utilizing 2.5D slicing for enhanced performance, showcasing promise for clinical applications.

## 1. Introduction

The aging process and certain neuropsychiatric diseases are often accompanied by a range of complex structural and functional brain changes. Recently, the magnetic resonance imaging (MRI) driven brain age paradigm has emerged as a promising biomarker for detecting variations across different diseases and health conditions (Gaser and Kalc et al., 2024). Research suggests that the deviation, known as brain age gap (BAG), between estimated brain age from MRI and actual age can serve as a widely accepted concept in disease diagnosis (Schmaal and Veltman et al., 2016; Cole and Franke, 2017; Chung and Addington et al., 2018; Cole and Marioni et al., 2019; Bashyam and Erus et al., 2020; Mishra and Beheshti et al., 2023), detecting abnormalities in the human brain and revealing associations with lifestyle habits like smoking (Linli and Feng et al., 2022) and drinking (Angebrandt and Abulseoud et al., 2022), as well as life satisfaction and resilience (Sone and Beheshti et al., 2022). The key in brain age estimation tasks is to train a brain age regression model using MRI data from healthy individuals, aiming for the most accurate result. Recently, state-of-the-art deep learning architectures have been applied to brain age estimation tasks, resulting in high performance measured by mean absolute error (MAE) of brain age estimation (Peng and Gong et al., 2021; He and Feng et al., 2022; Wood and Kafiabadi et al., 2022). However, there are still numerous factors influencing the clinical application of these models especially in the credibility of the estimated brain age results (Tanveer and Ganaie et al., 2023).

The use of BAG for disease diagnosis assumes that the examined brain follows a typical developmental pattern, with morphological changes caused by diseases potentially reflecting a deviation from the typical aging process. This hypothesis is particularly applicable to age-related neurodegenerative diseases, such as Alzheimer’s disease, where the brain may show signs of accelerated aging. However, for other conditions like bipolar disorder and mild cognitive impairment, the brain may instead exhibit a delayed or altered aging process, deviating from the typical trajectory in different ways (Franke and Gaser, 2012; Nenadić and Dietzek et al., 2017). Moreover, the hypothesis of deviations from the typical aging trajectory may not be universally applicable to the entire brain (Gutierrez Becker and Klein et al., 2018). If a model cannot provide confidence in the estimated results, it poses a crisis for the model’s clinical application. Uncertainty estimation techniques aim to provide confidence in estimated results, enhancing users’ trust in model outputs (Kim and Khanna et al., 2016). Introducing uncertainty estimation in brain age estimation allows measuring the deviation of a new subject from observed results, eliminating the need for implicit assumptions of accelerated aging process (Gutierrez Becker and Klein et al., 2018).

Uncertainty prediction can be seen as an interpretability method aimed at quantifying the uncertainty of model predictions rather than directly explaining the internal decision-making process. Unlike traditional interpretability methods such as visualization and feature importance analysis (Ribeiro and Singh et al., 2016; Samek and Wiegand et al., 2017), uncertainty prediction provides a confidence assessment of prediction results, which is crucial for understanding and trusting model performance in real-world applications, such as medical diagnostics (Begoli and Bhattacharya et al., 2019) and financial prediction (Blasco and Sánchez et al., 2024). By highlighting areas of high uncertainty, this approach not only helps explain model behavior but also offers insights for model improvement and validation, enhancing the robustness and reliability of AI systems (Abdar and Pourpanah et al., 2021; Gawlikowski and Tassi et al., 2023).

Uncertainty is typically classified into aleatory uncertainty and epistemic uncertainty (Kiureghian and Ditlevsen, 2009). In the task of estimating brain age, aleatory uncertainty often arises from individual differences in the human brain and the randomness in data collection (Hahn and Ernsting et al., 2022). Increasing the sample size does not necessarily reduce aleatory uncertainty. Epistemic uncertainty refers to uncertainty caused by insufficient model knowledge. This type of uncertainty can be mitigated by augmenting training data (Gawlikowski and Tassi et al., 2023). In brain age estimation, both types of uncertainties should be considered simultaneously. While traditional methods for uncertainty estimation, such as linear relevance vector regression (RVR) (Baecker and Dafflon et al., 2021) and Gaussian process regressors (GPR) (Gutierrez Becker and Klein et al., 2018), are capable of estimating both types of uncertainties, their time complexity becomes prohibitively high, especially when handling high-dimensional data inputs like medical images. Neural networks have been employed to address this challenge. In the field of medical imaging, two commonly used uncertainty methods based on deep learning are Deep Ensemble (Lakshminarayanan and Pritzel et al., 2017), an ensemble-based method, and Monte Carlo Dropout (MCDO) (Gal and Ghahramani, 2016), a Bayesian method (Kurz and Hauser et al., 2022). MCDO only requires activating the dropout layer used during training in the inference phase to obtain uncertainty estimates through multiple inferences. Deep Ensemble requires training multiple models to estimate uncertainty. While the training cost of Deep Ensemble is higher than that of MCDO, it provides better uncertainty estimation performance.

There has been relatively limited research on uncertainty in the task of brain age estimation. Becker et al. (Gutierrez Becker and Klein et al., 2018) used the GPR method for brain age estimation of MRI images, introducing GPR as an uncertainty estimation into neuropathology for the first time. Palma et al.(Palma and Tavakoli et al., 2020) used Quantile Regression (QR) for uncertainty estimation, but only considered aleatory uncertainty without considering epistemic uncertainty estimation. Hahn et al. (Hahn and Ernsting et al., 2022) proposed the Monte Carlo dropout composite quantile regression (MCCQR) based on neural networks. This method achieved a lower MAE while estimating aleatory uncertainty and epistemic uncertainty simultaneously. Yet, it is not a deep learning method itself, and only traditional methods were considered in the comparison. Shi et al. (Shi and Yan et al., 2020) used the Deep Ensemble method based on attention architecture to compare the performance of 2D slices, 2D multi-images, and 3D images in the estimation of fetal brain age and uncertainty, but their research is limited to the field of newborn infants. Hepp et al.(Hepp and Blum et al., 2021) used a heteroscedastic noise model for uncertainty estimation of 3D brain image data and provided a visual explanation of uncertainty. However, this study did not compare with other uncertainty methods and lacked estimation of epistemic uncertainty. In summary, there is a lack of deep learning-based uncertainty estimation in the field, particularly in simultaneously estimating aleatoric uncertainty and epistemic uncertainty.

One additional challenge in brain age estimation using deep learning is the issue of computational complexity. To fully leverage volumetric information, brain age estimation models based on 3D Convolutional Neural Networks are extensively utilized (Hwang and Yeon et al., 2021; Wood and Kafiabadi et al., 2022). Only a small portion of brain age models in the estimation task relies on 2D slices, primarily for the purpose of reducing computational complexity (Tanveer and Ganaie et al., 2023).The 2.5D slice technique, widely utilized in medical image detection(Kim and Lee et al., 2020) and segmentation (Li and Liao et al., 2022), involves capturing slices from axial, coronal, and sagittal planes, along with two additional oblique planes (45° and −45°) for enhanced analysis (Setio and Ciompi et al., 2016). Three vertically aligned plane images, along with additional oblique sectional planes, are input into distinct channels for training a 2.5D neural network model. Comparatively, the 2.5D model, as opposed to the 2D baseline model, can harness more dimensional information. In medical imaging tasks, the 2.5D model achieves superior performance without increasing the model parameters (Kim and Lee et al., 2020). As far as we know, 2.5D slices have not yet been introduced into the field of brain age estimation.

The final challenge lies in the measurement of uncertainty. The existing literature lacks consensus on the optimal measurement of uncertainty estimation quality, especially regarding uncertainty in regression problems. Hahn et al.(Hahn and Ernsting et al., 2022) utilized the Prediction Interval Coverage Probability (PICP) as a quantification metric for uncertainty. When PICP approaches a pre-defined confidence level, it indicates that the uncertainty of predictions has been correctly estimated and represented. Additionally, Negative Log Likelihood (NLL) is a commonly used metric in assessing uncertainty quality. NLL is the negative value of the Log Likelihood function, typically used to measure the model’s fit to observed data. A higher Log Likelihood value implies a higher probability of the model predicting observed data. Conversely, NLL, being the negative Log Likelihood function, is better when smaller. However, recent studies (Laves and Ihler et al., 2020) indicate that models typically exhibit lower errors on the training set, which can lead to an overconfidence in model predictions. Consequently, uncertainty is systematically underestimated when evaluated on the test set. In addition, estimating either epistemic uncertainty or aleatory uncertainty alone would result in a systematic underestimation of the overall uncertainty (Palma and Tavakoli et al., 2020). Both PICP and NLL metrics are subject to this systematic underestimation, thereby affecting the accurate comparison and evaluation of different models.

In recent years, Liu et al. (2020) have proposed the Spectral-normalized Neural Gaussian Process (SNGP), a single-network deterministic method, which only requires a simple modification of the neural network to enable the neural network to estimate uncertainty. While keeping the training and inference costs low, it is close to Deep Ensemble in terms of uncertainty estimation effect and surpasses other single-network deterministic methods. In this study, we will tackle the aforementioned challenges by employing the SNGP to the task of brain age estimation, enabling simultaneous estimation of both stochastic uncertainty and epistemic uncertainty. To comprehensively evaluate its performance, we will conduct a systematic comparison with other leading deep learning uncertainty estimation methods. Moreover, we will incorporate 2.5D slices into the brain age estimation process, aiming to enhance model performance while keeping the parameter count unchanged. Lastly, we will propose a suitable performance metric to effectively assess the quality of uncertainty estimation among different models. By employing these strategies, we aim to advance the field of brain age estimation and contribute to the development of more accurate and reliable uncertainty estimation methods.

## 2. Methods

### 2.1 Basic network architecture

We employed an architecture based on ResNeXt101 (Xie and Girshick et al., 2017), which represents an enhancement over ResNet (He and Zhang et al., 2016). The ResNeXt block is composed of a group of parallel residual modules. Each residual module consists of several branches, each having the same structure. These branches capture different feature subspaces and are merged together to generate the final output. Compared to ResNet, the main improvement in ResNeXt lies in the structure of the residual modules. In ResNet, each residual module contains only one branch. In ResNeXt, however, each residual module contains multiple branches, with the number of branches often referred to as the “cardinality”, determining the diversity within the module. By increasing the cardinality, ResNeXt can improve model performance without increasing the depth or width of the model. Recent applications of ResNeXt in medical image classification(Balnarsaiah and Nayak et al., 2023) and brain age prediction(Li and Hao et al., 2024) have demonstrated its effectiveness in capturing complex features. This makes ResNeXt particularly well-suited for our study, where it can handle multidimensional data effectively.

In this work, Deep ensemble utilizes two channels as outputs, μ(x) and ln(σ(x)), where μ(x) serves as the estimation for brain age, and ln(σ(x)) serves as the estimation for the aleatory uncertainty of input images. The variance of the brain age estimation results output by independently trained models is used as an estimation of epistemic uncertainty. MCDO employs a single-channel output for estimation, utilizing the variance of inference results obtained through multiple iterations to assess epistemic uncertainty. Meanwhile, we also consider the integration of MCDO with heteroscedastic noise networks——MCDO-HN, building upon the basis of utilizing the two-channel outputs of Deep ensemble. We conduct multiple inferences using MC-dropout, aiming to simultaneously capture epistemic uncertainty and aleatory uncertainty. SNGP, on the other hand, utilizes a specialized output layer——Gaussian process layer. Lastly, we also considered a simplified version of SNGP (without spectral normalization) for performance comparison. A summary of the methods used in the current work is presented in Table 1.

**Table 1.** Summary of all the methods used in the current work.

| Methods | Additional Regularization | Output Layer | Ensemble Training | Multi-pass Inference | The type of uncertainty of the estimate |
| --- | --- | --- | --- | --- | --- |
| SNGP | Spectral-normalization | GP | - | - | AU+EU |
| Deep ensemble | - | Dense | Yes | Yes | AU+EU |
| MCDO-HN | Dropout | Dense | - | Yes | AU+EU |
| GP | - | GP | - | - | AU+EU |
| MCDO | Dropout | Dense | - | Yes | EU |

*Note. AU refers to aleatory uncertainty, EU refers to epistemic uncertainty, and Multi-pass Inference refers to the necessity of multiple inferences to obtain an estimated result.*

### 2.2 Spectral-normalized Neural Gaussian Process

The Spectral-normalized Neural Gaussian Process (SNGP) is a method proposed by Liu et al.(Liu and Padhy et al., 2022) for quantifying uncertainty in deep learning. As shown in **Figure 1**, SNGP requires only simple modifications to neural networks to enable uncertainty estimation. Specifically, SNGP incorporates spectral normalization into the network, which helps maintain distance in the hidden space, enhancing distance-awareness capabilities. This ensures a correspondence between input and hidden space distances, crucial for improving the quality of quantifying model prediction uncertainty. The specific form of spectral normalization is as follows:

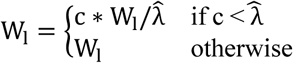

**Figure 1.**
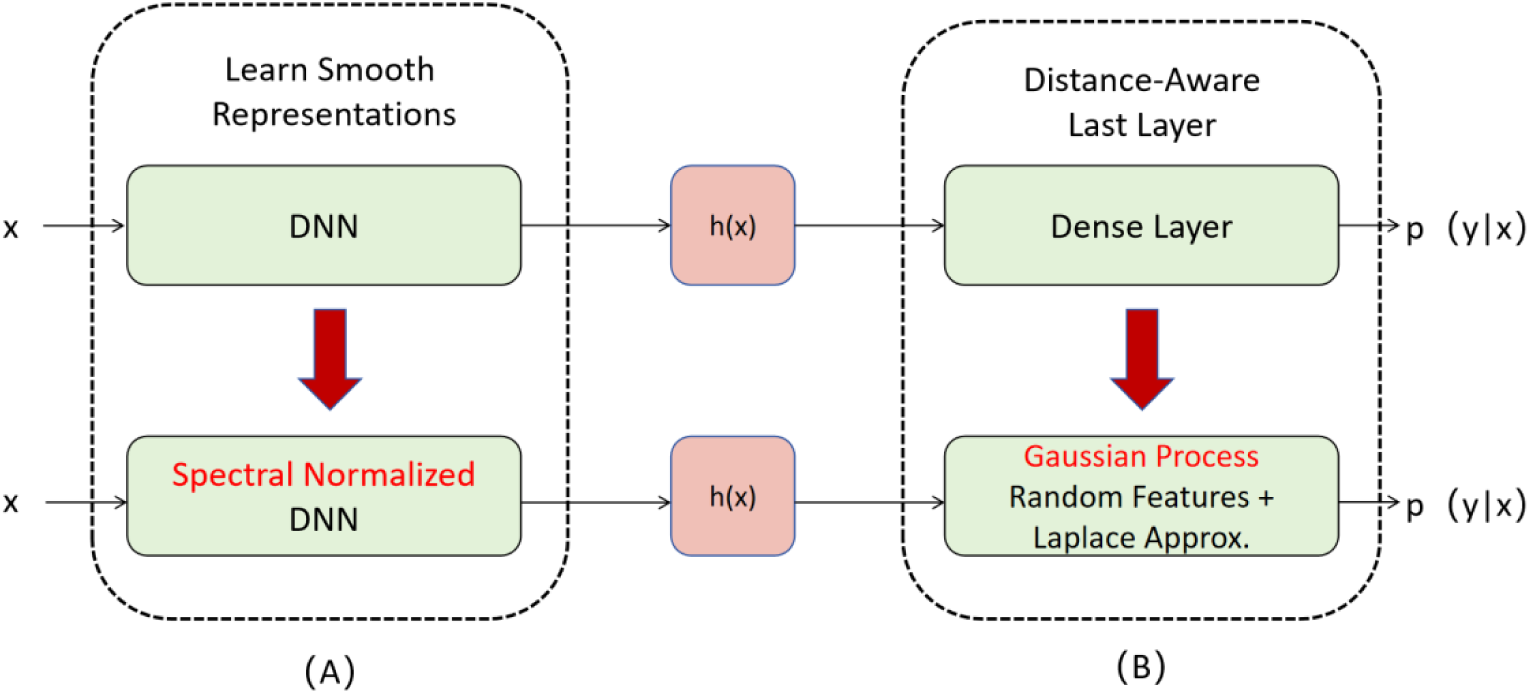
SNGP incorporates two modifications to deep neural networks: (A) the inclusion of spectral normalization layers in the hidden layers, and (B) replacing the final dense layer with a Gaussian process layer using random Fourier features and Laplace approximation.

W_l_ represents the weight matrix of the l-th layer, 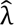 ≈ ‖W_l_‖_2_, where c is a hyperparameter controlling weight scaling. Furthermore, to endow SNGP with the capability of output distance awareness, the traditional dense layer is replaced by a Gaussian Process layer with a radial basis function (RBF) kernel. The output of the Gaussian Process layer is a multivariate normal distribution. As the distance between testing samples and the training dataset increases, the variance of the posterior distribution based on the RBF kernel Gaussian process also increases accordingly, reflecting higher uncertainty for input samples in unknown regions or far from the training data distribution. To ensure computational efficiency, SNGP employs Laplace-approximated random feature expansions to approximate the posterior distribution of the GP. This allows for scalable learning in closed form, with minimal modifications to the deterministic DNN training process, while effectively computing prediction uncertainty on a per-input basis.

Compared to mainstream uncertainty estimation methods, SNGP requires only single inference and single training, reducing training costs while also improving training efficiency. Besides, SNGP can provide high-quality uncertainty assessments. These advantages enable SNGP to perform exceptionally well in the field of medical imaging (Tabarisaadi and Khosravi et al., 2022; Ahmed and Abu Yousuf et al., 2023; Dorkenwald and Li et al., 2023). SNGP was initially applied in classification models, and its regression form was further extended in subsequent studies (Liu and Padhy et al., 2022) by the authors. This work employs its regression form.

This article adopts a 512*512 random Fourier feature (RFF) matrix to approximate the RBF kernel, utilizing the moving average method to update the covariance matrix, with a ridge factor of 1e-4 and a discount factor of 0.999, consistent with the recommendations of Liu et al. (Liu and Padhy et al., 2022) for other parameters. Spectral normalization layers are incorporated into each convolutional layer of ResNeXt101, and the final fully connected layer is replaced with a Gaussian process layer. For regression, the loss function of SNGP is as follows:

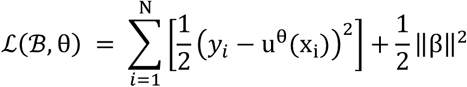

where N represents the total number of samples, x_i_ and denotes the input for each individual sample, u^θ^(x_i_) represents the estimation of brain age, β represents the weights of the final learnable output layer.

### 2.3 Other uncertainty estimation methods for comparison

MCDO, as proposed by Gal et al. (Gal and Ghahramani, 2016), simulates Bayesian inference by applying dropout during the inference phase. This technique involves performing multiple forward passes, with each pass randomly dropping a subset of neuron outputs, effectively sampling from various sub-models to approximate the prediction distribution and quantify uncertainty. For each input sample, MCDO computes the mean of outputs from these passes as the final prediction and uses the variance to measure uncertainty. The loss function of MCDO is as follows:

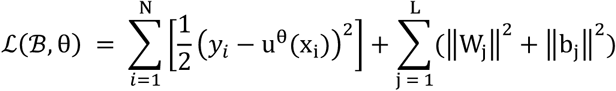

where u^θ^(x_i_) represents the estimation of brain age, represents the weights of the final learnable output layer.W_j_ represents the weights of the j-th layer,b_j_ represents the biases of the j-th layer.

MCDO employs standard loss functions, without requiring complex modifications to the network architecture. It is widely used in fields like medical image analysis (Hepp and Blum et al., 2021; Hahn and Ernsting et al., 2022; Ernsting and Winter et al., 2023), providing a straightforward and effective means to assess the confidence and reliability of model predictions.

Deep Ensemble, proposed by Lakshminarayanan et al. (Lakshminarayanan and Pritzel et al., 2017),enhances uncertainty estimation by training multiple neural networks independently. Each model in the ensemble has different initializations, and optionally, different subsets of the training data. The method computes the mean of the predictions from all models as the final output and uses the variance to quantify uncertainty. Deep ensemble has two outputs μ(x) and ln(σ(x)), with Negative Log-Likelihood (NLL) serving as the network’s loss function, as shown below:

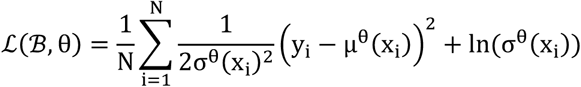

where the network estimate σ^θ^(x_i_)^2^ is considered as an estimate of aleatory uncertainty, μ^θ^(x_i_) represents the estimation of brain age. Deep ensemble independently trains M networks with randomly initialized parameters, estimating epistemic uncertainty through the variance of different network estimates. Consistent with prior research, we set M=5 in this study (Lakshminarayanan and Pritzel et al., 2017; Beluch and Genewein et al., 2018).

The Monte Carlo Dropout with heteroscedastic noise networks (MCDO-HN) also has two outputs and its loss function are consistent with Deep Ensemble. However, the method differs in how it estimates uncertainty. In the study of Hepp et al.(Hepp and Blum et al., 2021), this method was used as a brain age estimation from 3D brain images.

The Gaussian Process (GP) method, apart from not incorporating Spectral-normalized layers in the network, is essentially the same as SNGP, with its loss function remaining consistent with that of SNGP. In the study by Liu et al(Liu and Padhy et al., 2022). The GP method is introduced as a control method for comparing with SNGP.

### 2.4 Uncertainty estimation

MCDO, conversely, estimates epistemic uncertainty by randomly dropping neurons in the network and computing the variance of results through M inferences. The specific computation methods for estimates uncertainties and brain age estimation are as follows:

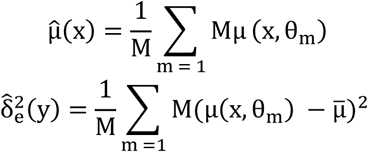

MCDO-HN estimates epistemic uncertainty based on this foundation, with the calculation method as follows:

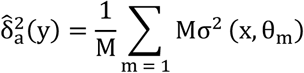

Ultimately, epistemic uncertainty and aleatory uncertainty jointly constitute the network’s prediction of uncertainty 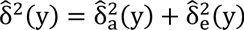. The Deep Ensemble method also uses the same computational approach, but the epistemic uncertainty of the Deep Ensemble is provided by five independently trained models. In the SNGP and GP, the variance of the output from the Gaussian process layer is directly employed as an estimate of uncertainty.

## 3. Experiments

### 3.1 Analytical Overview

Figure 2 outlines the analytical pipeline used in this study. We firstly collected data from 11 publicly available T1-weighted brain sMRI datasets (N=6628) and employed the CAT12 toolbox for data preprocessing. Furthermore, a 2.5D slicing method was applied to slice the sMRI data. Five-fold cross-validation was conducted on these 11 datasets to compare the performance of SNGP with other mainstream methods. Performance estimation was based on MAE, while uncertainty performance was evaluated using Pearson correlation coefficients. Additionally, the Neurocognitive aging dataset (N=301) was incorporated to assess the generalization performance of the models. Lastly, controlled experiments were conducted to compare patients versus non-patients, Asian versus non-Asian populations, 2.5D versus 2D methods,DenseNet Backbone versus ResNeXt Backbone,and Site effect correction versus without Size effect correction, thus enhancing the assessment of the models’ performance.

**Figure 2.**
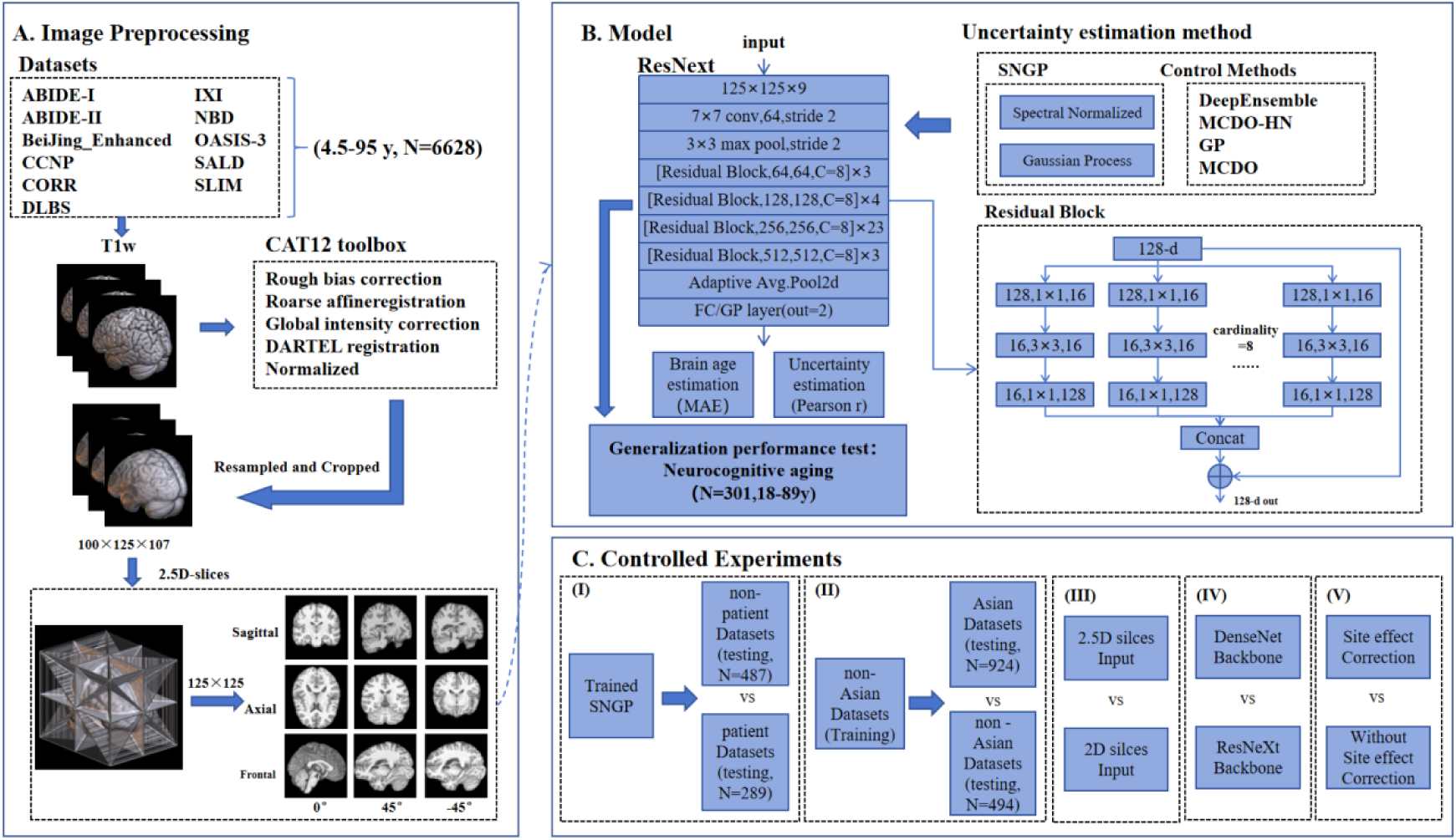
Flow diagram of the analysis approach used in the study, including (A) image preprocessing, (B) model training, and (C) five controlled experiments.

### 3.1 Datasets

Similar to the majority of brain age estimation models, this study employs a regression model trained on T1-weighted MRI images from healthy individuals to predict brain age. We collected 6628 samples from 12 publicly available datasets containing healthy brain T1-weighted MRI scans (refer to **Table 2**). These scans cover ages ranging from 4.5 to 97 years, with a nearly equal gender distribution and a balanced representation across various age groups. Eleven datasets were utilized for cross-validation, while one dataset was reserved for assessing the model’s generalization ability.

**Table 2.** Demographic Information of Brain Age Estimation Datasets Used in This Study.

| Dataset | N<sub>samples</sub> | Age range | Age(mean±standard deviation) | Gender (female/male) |
| --- | --- | --- | --- | --- |
| ABIDE-I<br>(Di Martino and Yan et al., 2014) | 570 | 6.47-56.2 | 17.10±7.73 | 472/98 |
| ABIDE-II<br>(Di Martino and O Connor et al., 2017) | 557 | 5.887-64 | 15.26±9.51 | 382/175 |
| BeiJing_Enhanced<sup>1</sup> | 180 | 17-28 | 21.22 ± 1.93 | 73/107 |
| CCNP(Liu and Wang et al., 2021) | 459 | 4.5-18.9 | 10.43 ± 2.90 | 241/218 |
| CORR(Zuo and Anderson et al., 2014) | 1527 | 6-88 | 25.94 ± 15.28 | 765/762 |
| DLBS(Park and Carp et al., 2012) | 315 | 20.57-89.11 | 54.62 ± 20.06 | 117/198 |
| IXI<sup>2</sup> | 563 | 20-86.31 | 48.65 ± 16.46 | 250/313 |
| NBD(Nastase and Liu et al., 2021) | 334 | 18-53 | 21.93 ± 4.67 | 136/198 |
| OASIS-3<br>(LaMontagne and Benzinger et al., 2019) | 754 | 42.5-95 | 66.07 ± 9.00 | 304/450 |
| SALD(Wei and Zhuang et al., 2018) | 494 | 19-80 | 45.18 ± 17.43 | 187/307 |
| SLIM(Liu and Wei et al., 2017) | 574 | 17-27 | 20.09 ± 1.28 | 254/320 |
| *Neurocognitive aging | 301 | 18-89 | 40.94 ± 23.05 | 169/132 |
| Overall | 6628 | 4.5-95 | 31.93 ± 21.45 | 3127/3146 |

<sup>1</sup> https://fcon_1000.projects.nitrc.org/indi/retro/BeijingEnhanced.html
<sup>2</sup> https://brain-development.org/ixi-dataset/
\*Datasets employed for assessing generalizability were obtained from OpenNeuro (https://openneuro.org/datasets/ds003592).

### 3.2 Pre-processing

T1-weighted images were preprocessed with CAT12 toolbox (Gaser and Dahnke et al., 2024) with default settings in SPM12(https://www.fil.ion.ucl.ac.uk/spm/software/). The steps include rough bias correction, coarse affine registration, global intensity correction, and segmentation. Then DARTEL registration were run with MNI 152 template (Ashburner, 2007). The bias, noise, global intensity corrected and normalized T1 images were used in the following analysis. All images were resampled and cropped to an isotropic spatial resolution of 1.5×1.5×1.5 mm^3^ and an image size of 100×125×107 mm^3^.

In this work, we employed a 2.5D slicing method (Setio and Ciompi et al., 2016) which better utilizes spatial information compared to 2D slicing. As shown in Figure 3, the 2.5D slicing acquires medical slices from axial, coronal, and sagittal directions, and rotates each slice by ±45 degrees to obtain two oblique slices for each direction. The nine slices, after cropping the black borders, were standardized to a size of 125×125 and input into the channels. Simultaneously, for comparison, we introduced 2D slicing. To match the channel numbers between 2D and 2.5D, we considered nine transverse slices, with four slices above the central slice and four slices below the central slice. The size of the 2D slices was 100×125.

**Figure 3.**
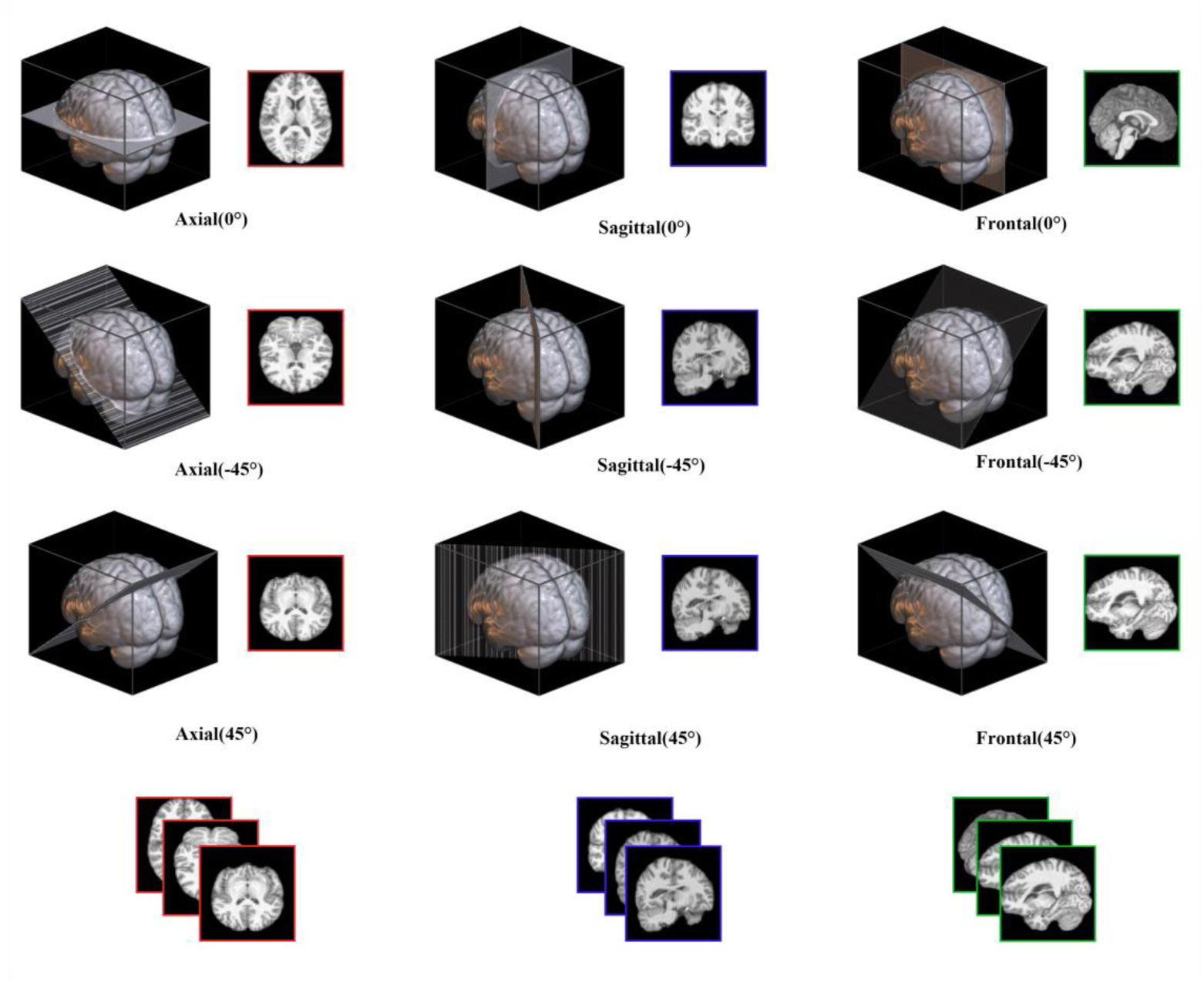
The 2.5D MRI images consist of slices in the axial, coronal, and sagittal planes, with two additional oblique intersecting planes (at 45° and −45°) for each plane.

### 3.3 Experimental details

We employed 5-fold cross-validation, utilizing the Adam optimizer with an initial learning rate of 1e-4, a batch size of 64, and conducted training for 600 epochs. PyTorch was selected as the deep learning framework(Paszke and Gross et al., 2019), and computations were executed on an RTX A4000 GPU with 16GB of memory.

### 3.4 Uncertainty quantification

The existing uncertainty quantification methods in academia each have their own limitations, and there is no consensus within the scholarly community on how to measure regression uncertainty. A good uncertainty estimation should reflect the accuracy of model predictions. That is, when the model provides greater uncertainty, prediction errors should also be larger, and vice versa. Therefore, we propose an index to compare the uncertainty estimation performance, calculated by the Pearson correlation coefficient between Mean Absolute Error (MAE) and the standard deviation of model estimates (square root of uncertainty estimation results) to compare the uncertainty estimation performance of different models. The Pearson correlation coefficient can be dimensionless, unaffected by systematic underestimation of uncertainty, and facilitates direct comparison of different estimation methods (some of which only consider epistemic uncertainty).

### 3.5 Controlled Experiments

As previously mentioned, the differences in BAG may arise not only from intrinsic changes in the brain itself but also from the high uncertainty of the models. Following the definition by Hahn et al.(Hahn and Ernsting et al., 2022), this study incorporates uncertainty into the consideration of BAG, presenting the concept of uncertainty-adjusted BAG through the following formula:

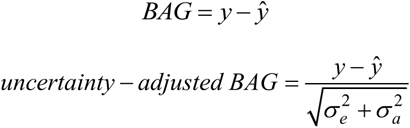

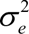 and 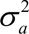 represent cognitive uncertainty and stochastic uncertainty, respectively.

This study utilized a pre-trained SNGP model and the ADHD200 (N=776) dataset for testing. ADHD, Attention Deficit and Hyperactivity Disorder, affects the development of the brain(Castellanos and Lee et al., 2002; Shaw and Eckstrand et al., 2007), thereby exhibiting differences in BAG compared to the normal control group. BAG and uncertainty-adjusted BAG were categorized by disease status for t-tests. The patient data consisted of N=289 samples, while the non-patient data consisted of N=487 samples.

Secondly, we investigated the Out of Distribution (OOD) detection capability of SNGP, and BAG exhibits significant differences across different races(Dempsey and Deardorff et al., 2023). If a model is trained on one race, its estimation performance on other races will be comparatively poor(Hahn and Ernsting et al., 2022). This requires the model to exhibit different levels of uncertainty in its output when the distribution of input samples in the test set differs from that of the training set. This study trained the model using non-Asian datasets (ABIDE I, ABIDE II, CORR, DLBS, IXI, NBD, OASIS-3), with 20% randomly selected as the test set (N = 924). Additionally, the SALD dataset (N = 494) was selected as an Asian dataset, and t-tests were employed to assess the disparity in uncertainty between Asian and non-Asian dataset outputs.

Thirdly, we conducted an experiment where we replaced the SNGP model’s backbone with DenseNet (Gao Huang, 2017), specifically the DenseNet121 version. We used the same five-fold cross-validation to ensure consistency in the evaluation process. This allowed us to evaluate the robustness and generalization capabilities of our approach using a different network architecture and provided a more detailed comparison of performance between the two models. DenseNet features dense connections where each layer receives input from all preceding layers. This design mitigates the vanishing gradient problem and promotes feature reuse for efficient learning. In medical imaging, DenseNet has been applied to disease detection (Chauhan and Palivela et al., 2021), image segmentation (Gottapu and Dagli, 2018), and enhancement (Tong and Gou et al., 2019), significantly improving diagnostic accuracy and image quality (Zhou and Ye et al., 2022).DenseNet and ResNeXt architectures demonstrate comparable performance metrics in predictive tasks (Bianco and Cadene et al., 2018), we chose to use DenseNet not to compare its performance with ResNeXt, but to demonstrate that SNGP can be broadly applied to various neural network architectures.Recent studies have shown that DenseNet performs well on brain age estimation tasks (Wood and Kafiabadi et al., 2022).

Fourth, site effects are an issue that warrants attention (Mishra and Beheshti et al., 2023). However, recent studies indicate that not correcting for these effects might actually yield better results, with higher accuracy in age prediction observed when site correction is not applied(Yu and Cui et al., 2024). Nevertheless, to evaluate and confirm potential biases and non-biological influences of acquisition site effects on our model, we performed the ComBat harmonization procedure (Johnson and Li et al., 2007) on the preprocessed images using the DPABI Harmonization module (Wang and Chen et al., 2023). Age and sex ere included as covariates. The adjusted data was then used to retrain the SNGP model using five-fold cross-validation.

Finally, to evaluate the performance enhancement achieved by incorporating 2.5D slices, we retrained the SNGP model with minimal modifications to the network architecture. Utilizing the same five-fold cross-validation approach and the 2D slice method ensured a fair comparison, allowing us to accurately assess the impact of the 2.5D slices on model performance. The backbone network employed in our experiments was ResNext101.

Table 3 provides a concise summary of the controlled experiments conducted in this study, highlighting the uncertainty estimation methods, backbone architectures, and datasets used for both training and testing.

**Table 3.**
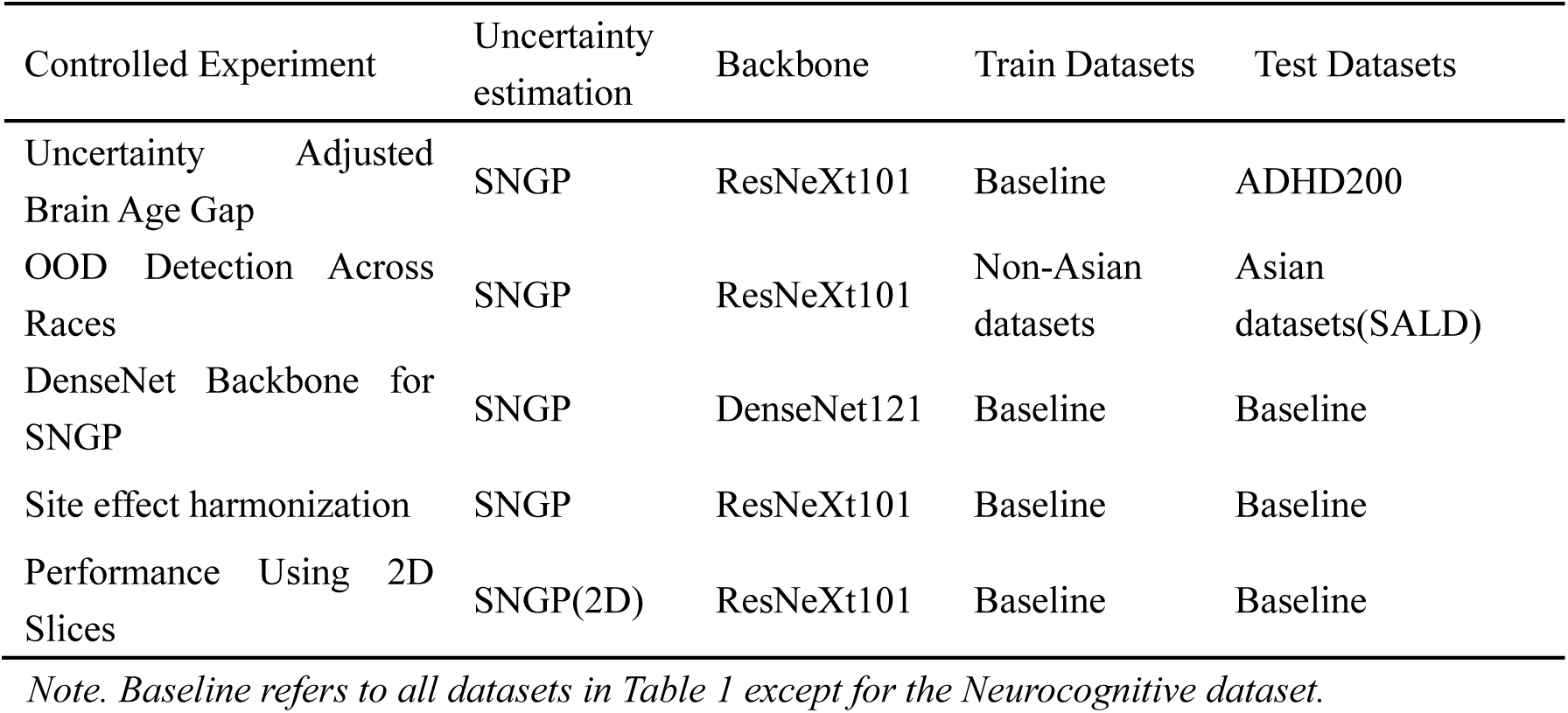
Summary of all the Controlled Experiments used in the current work.

## 4. Result

### 4.1 Model performance

We compared the SNGP model with two commonly used deep learning baseline uncertainty estimation methods, namely DeepEnsemble and MCDO. In addition, this paper introduces a reduced version of SNGP called DNN-GP (which removes spectral normalization from SNGP) and MCDO-HN (combining MCDO with heteroscedastic noise networks) for comparison. The predictive performance of the models was evaluated using the MAE.

We repeatedly trained our models using five-fold cross-validation on 11 publicly available datasets. As shown in **Table 4**, the MAE for SNGP was 2.95 years (SD=0.14), while for other methods, it ranged from 2.80 to 2.63. The performance of SNGP, possibly influenced by modifications such as spectral normalization and Gaussian process output layers, is affected by the complexity of the network, leading to suboptimal predictive performance compared to other methods, consistent with previous research findings (Liu and Lin et al., 2020). Nevertheless, SNGP’s predictive performance remains competitive compared to mainstream deep learning models for brain age prediction. MCDO performed the best in terms of MAE, reaching 2.63 years (SD=0.06), and its modifications to the network are relatively simple.

**Table 4.** Model performance comparison.

| Models | Brain age estimation performance<br>(MAE) |  | Uncertainty estimation performance<br>(Pearson correlation R) |  |
| --- | --- | --- | --- | --- |
|  | 5-Fold CV | Neurocognitive aging | 5-Fold CV | Neurocognitive aging |
| SNGP | 2.95(0.14) | 3.43 | 0.554 | 0.460 |
| Deep ensemble | 2.80(0.14) | 3.27 | 0.535 | 0.317 |
| MCDO-HN | 2.90(0.06) | 3.2 | 0.446 | 0.368 |
| GP | 2.81(0.13) | 3.11 | 0.324 | 0.214 |
| MCDO | 2.63(0.11) | 3.07 | 0.383 | 0.334 |
| *SNGP<br>(DenseNet) | 2.85(0.06) | 3.24 | 0.598 | 0.481 |
| *SNGP(2D) | 3.04(0.05) | 3.62 | 0.542 | 0.381 |
| *SNGP(site effects<br>harmonization) | 3.02(0.08) | 3.45 | 0.474 | 0.421 |

*Note. For cross-validation, SD across folds is given in parentheses. \* stands for Controlled Experiments*

Although five-fold cross-validation is sufficient to demonstrate the model’s generalization ability, the model may be influenced by different data collection protocols or sample characteristics. Therefore, we introduced the Neurocognitive aging dataset as an independent validation set. The MAE on Neurocognitive aging dataset generally higher than the results of five-fold cross-validation, but the ranking of model performance is similar to the cross-validation results. On Neurocognitive aging dataset, the MAE for SNGP is 3.43 years, while MCDO reaches 3.07 years, and other methods range from 3.11 to 3.27 years. The comparative visualization of the results from various methods is presented in **Figure 4**. Previous studies have reported biases in BAG across different age groups.(Peng and Gong et al., 2021) To further investigate this aspect, we analyzed the correlation between BAG and chronological age using the SNGP model. As shown in **Figure 5**, the results demonstrate a relatively weak correlation, suggesting that the BAG estimated by SNGP is less influenced by age compared to other models.

**Figure 4.**
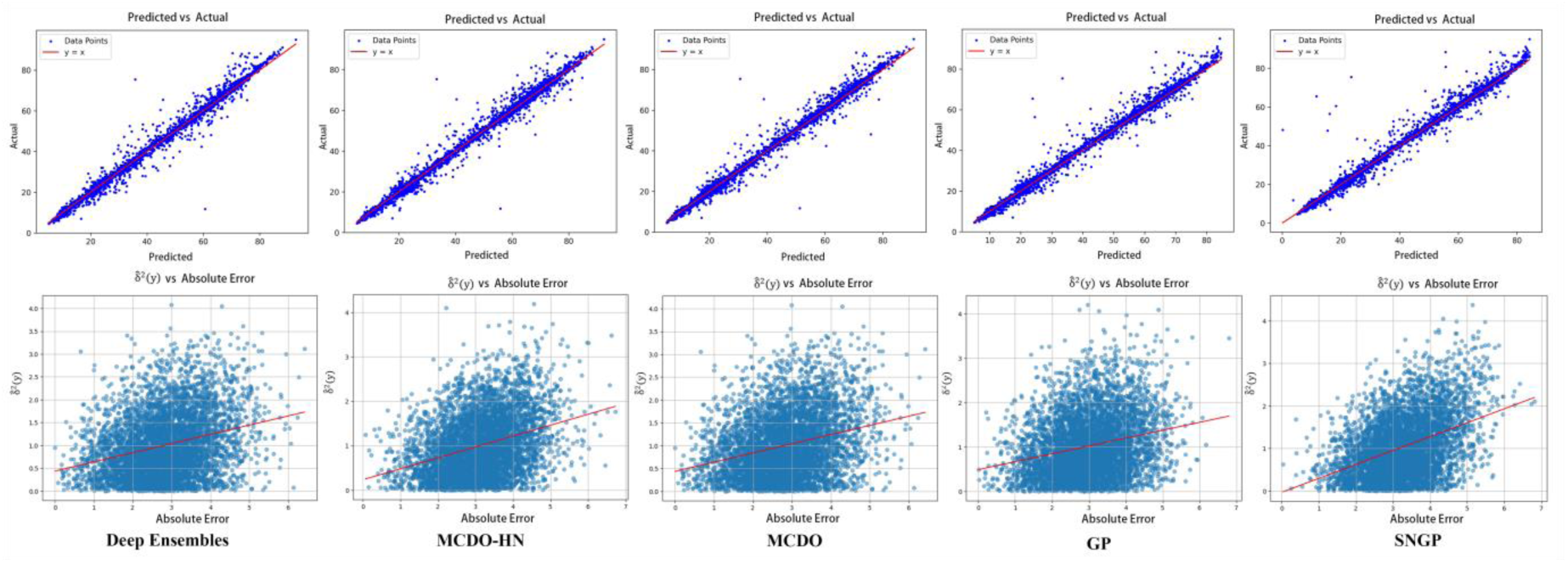
The top panel of figure illustrates the correlation between the predicted brain ages and the actual brain ages for different methods, in which the x-axis represents the predicted age, while the y-axis denotes the actual age. The bottom panel depicts the relationship between the predicted uncertainty of each method and the absolute errors in brain age predictions, in which the x-axis corresponds to the absolute error, and the y-axis represents the estimated uncertainty.

**Figure 5.**
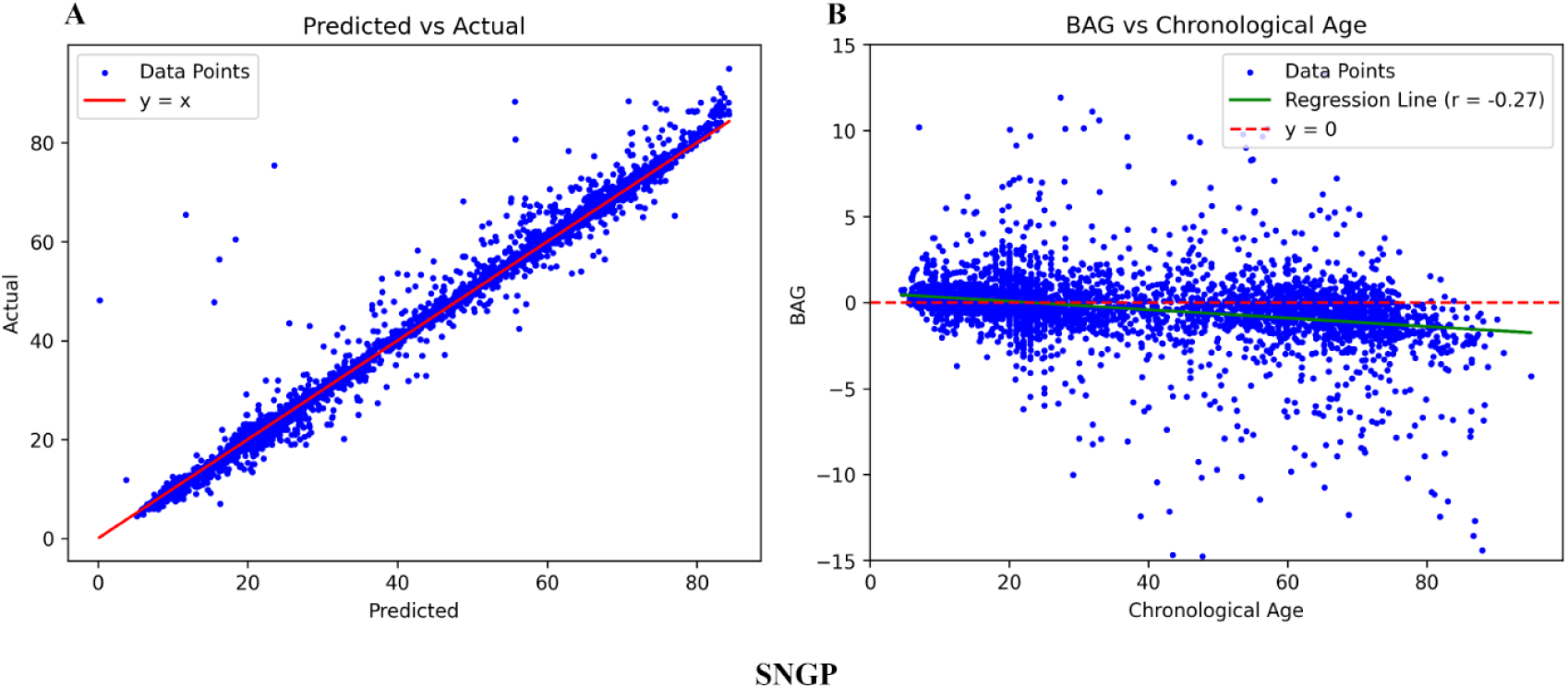
(A) presents the correlation between predicted and actual brain ages using the SNGP model, while (B) illustrates the BAG versus chronological age, showing a regression slope of r = - 0.27.

### 4.2 Uncertainty estimation performance

In **Table 4**, the uncertainty quality predicted by SNGP is the highest, with r = 0.554, while DeepEnsemble also achieved a high correlation of r = 0.535. The uncertainty estimation quality of SNGP and DeepEnsemble is roughly equivalent, but SNGP is a single inference model, with training and inference costs much lower than Deep Ensemble. The Pearson correlation coefficient of GP is only r = 0.365, indicating that the uncertainty estimation quality of SNGP is much higher than that of GP. This suggests that spectral normalization ensures the network’s ability to discern distances in the hidden space, which is crucial for high-quality uncertainty estimation. MCDO-HN incorporates the ability to predict stochastic uncertainty on top of MCDO, resulting in a significant improvement in uncertainty quality. This underscores the necessity of considering both aleatory uncertainty and epistemic uncertainty in uncertainty estimation.

In terms of generalization performance, SNGP also demonstrates the best generalization ability, maintaining r = 0.46 on the Neurocognitive aging dataset. Both Deep Ensemble (r = 0.317) and GP (r = 0.214) show substantial declines in performance, while MCDO-HN exhibits a smaller decrease in performance. Overall, SNGP achieves the highest uncertainty estimation quality and the best generalization ability.

### 4.3 Results of three Controlled Experiments

#### 4.3.1 Uncertainty Adjusted BAG

On the inputs of patients and non-patients from the ADHD dataset, the results indicated that the t-test for BAG yielded (t = 2.62, p = 0.034, Cohen’s d = 0.168), with the model passing the test at a significance level of 5%. The t-test for uncertainty-adjusted BAG yielded (t = 3.131, p = 0.002, Cohen’s d = 0.233), passing the test at a significance level of 1%, with a 38.6% increase in effect size (Cohen’s d). The 38% improvement was calculated based on the Cohen’s d values, using the formula 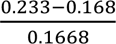 × 100% = 38.6%, where 0.233 and 0.168 represent the effect sizes for uncertainty-adjusted BAG and BAG, respectively. This may suggest the need to consider the impact of uncertainty in brain age estimation tasks.

#### 4.3.2 OOD Detection Across Races

On the inputs from Asian and non-Asian individuals, the uncertainty of the model’s output was tested using t-tests based on Asian and non-Asian datasets, revealing significant differences in uncertainty (t = 2.701, p = 0.007). The results demonstrate that the model possesses a certain degree of ODD capability.

#### 4.3.3 DenseNet Backbone for SNGP

As shown in **Table 4**, the performance of the SNGP method varies with different backbones. The five-fold cross-validation performance of DenseNet is slightly better than that of ResNeXt, with a Mean Absolute Error (MAE) of 2.85 and an uncertainty prediction performance of 0.598. The model also performs well on the independent validation set, achieving a MAE of 3.24 and a Pearson correlation coefficient of 0.481. This advantage may be attributed to DenseNet’s feature reuse capability, which allows gradients to propagate more easily from deeper layers to shallower layers, thereby mitigating the vanishing gradient problem. DenseNet’s excellent performance in the medical imaging field aligns with the expectations of this study. This also demonstrates the inherent adaptability of the SNGP method, proving its applicability across various network architectures.

#### 4.3.4 Site effect harmonization

In **Table 4**, after correcting for site effects, the mean absolute error (MAE) in five-fold cross-validation reached 3.02, which is worse than the original 2.95, with a correlation coefficient of 0.474. In the independent validation set, the MAE was 3.45, with a correlation coefficient of 0.421. The performance of the model declined after correcting for site effects, consistent with the conclusions presented in previous studies (Yu and Cui et al., 2024).

#### 4.3.5 Performance Using 2D Slices

Finally, the performance of the SNGP method significantly declines when using the ResNeXt backbone with 2D slices. The Mean Absolute Error (MAE) increases to 3.04, and the Pearson correlation coefficient drops to 0.542. This performance degradation is even more pronounced in the independent validation set, with a MAE of 3.62 and a Pearson correlation coefficient of 0.381. These results indicate that the 2.5D slices method improves the model’s performance without increasing network parameters or computational burden.

## 5. Discussion

To our best knowledge, the current work firstly introduces the SNGP method into the field of deep learning brain age estimation. SNGP only requires simple modifications to the network to enable it to estimate uncertainty, and it only requires single training and inference. The model demonstrates the best uncertainty estimation performance on 11 public datasets and strong generalization capabilities on an independent validation sample. This indicates that SNGP can adapt well to the task of brain age uncertainty estimation. In deep learning networks, uncertainty estimation models need to incorporate weight normalization steps to improve distance-aware output capability. In practice, removing spectral normalization from the SNGP network leads to a decrease in uncertainty estimation performance, thus spectral normalization is necessary. Additionally, we replaced the backbone of SNGP from ResNeXt to a similarly performing DenseNet and conducted a 5-fold cross-validation. This change resulted in a certain performance improvement, which also demonstrates that SNGP can adapt to various network architectures, exhibiting good generalization performance.

Existing deep learning uncertainty estimation methods either have low-quality uncertainty estimation, high training and inference costs, or only provide aleatory or epistemic uncertainty estimates (Gutierrez Becker and Klein et al., 2018; Hepp and Blum et al., 2021; Hahn and Ernsting et al., 2022). In contrast, SNGP offers single training and single inference, while allowing for the estimation of both aleatory and epistemic uncertainties. This approach not only reduces computational and inferential costs but also provides more accurate estimates of uncertainty, facilitating the practical application of brain age estimation models in clinical settings.

Building upon this, this paper uses the ADHD200 dataset to detect the brain age difference between patients and non-patients and corrects the BAG considering uncertainty. Uncertainty-adjusted BAG enhances the ability to detect changes in brain development in adolescents with ADHD. Therefore, considering uncertainty estimation in brain age estimation is crucial. For example, in clinical settings, directly using uncertainty as a BAG for calibration, or introducing medical imaging experts to reassess the results if the model outputs uncertainty exceeding the confidence interval, is necessary. Besides, SNGP demonstrates the ability to detect ODD between different races, as there are significant differences in BAG between different races (Dempsey and Deardorff et al., 2023), and having ODD capability is also essential for the clinical application of brain age estimation models.

Additionally, the introduction of 2.5D slices improves the estimation performance of the network. The implementation of 2.5D slices is simple and can be easily applied to other 2D networks. Subsequent research can employ 2.5D slices in other 2D networks to enhance their performance. After correcting for site effects, the model’s performance declined, which aligns with a recent study (Yu and Cui et al., 2024) suggesting that it may be better not to correct for site effects. Future research might not need to consider correcting for site effects.

There are several limitations in this study. Firstly, as current spectral normalization techniques cannot precisely control the true spectrum regularization of convolutional kernels, and other regularization mechanisms (such as BatchNorm) may reassign the spectrum regularization of a layer in unexpected ways (Gouk and Frank et al., 2021), thereby affecting the estimation performance of SNGP. Subsequent research may need to focus on reducing conflicts between spectral normalization and other regularization methods. Secondly, SNGP undergoes the most extensive modifications to the network and causes greater estimation performance loss compared to other methods. Subsequent research needs to further reduce the loss in estimation performance. But SNGP is not an independent brain age estimation method. SNGP can be applied to any state-of-the-art deep learning brain age estimation method, such as SFCN (Peng and Gong et al., 2021) or Global-local transformer (He and Grant et al., 2022), to enable these advanced networks to estimate uncertainty.

Currently, there is still controversy over how to evaluate the quality of uncertainty estimation in regression tasks (Sluijterman and Cator et al., 2024), especially regarding the comparison between aleatory and epistemic uncertainties. The method proposed in this paper, which uses the Pearson correlation coefficient between MAE and the square root of uncertainty, represents just one approach. This method can prevent the model from underestimating uncertainty due to too low MAE in the training set and facilitates the comparison of different uncertainty estimation methods. Empirical evidence shows that using the Pearson correlation coefficient as an evaluation of uncertainty quality aligns with theoretical expectations and is consistent with SNGP’s performance in other tasks (Liu and Padhy et al., 2022).

## Data Availability

All data produced in the present study are available upon reasonable request to the authors.

